# COVID-19 mRNA vaccine is not detected in human milk

**DOI:** 10.1101/2021.03.05.21252998

**Authors:** Yarden Golan, Mary Prahl, Arianna Cassidy, Christine Y. Lin, Nadav Ahituv, Valerie J. Flaherman, Stephanie L. Gaw

**Affiliations:** Department of Bioengineering and Therapeutic Sciences, University of California, San Francisco; Institute for Human Genetics, University of California, San Francisco; Department of Pediatrics, University of California, San Francisco; Division of Pediatric Infectious Diseases and Global Health, University of California, San Francisco; Division of Maternal-Fetal Medicine, Department of Obstetrics, Gynecology, and Reproductive Sciences, University of California San Francisco

## Abstract

Several countries have recently approved the use of mRNA vaccines against COVID-19 under an emergency use authorization. However, no pregnant or lactating individuals were included in the Phase 3 clinical trials of these vaccines despite belonging to a group at high risk for severe complications of COVID-19 infection. We show here that the mRNA from anti-COVID BNT162b2 (Pfizer) and mRNA-1273 (Moderna) vaccines is not detected in human breast milk samples collected 4-48 hours post-vaccine. These results strengthen the recommendation of ABM and WHO that lactating individuals who receive the anti-COVID-19 mRNA-based vaccine should continue to breastfeed their infants uninterrupted.

## Introduction

Several countries have recently approved the use of mRNA vaccines against COVID-19 under an emergency use authorization ^1^. However, no pregnant or lactating individuals were included in the Phase 3 clinical trials of these vaccines despite belonging to a group at high risk for severe complications of COVID-19 infection ^2,3^. As a result, there are no clinical data regarding the safety or efficacy of the vaccine in these populations. The World Health Organization (WHO) recommends breastfeeding people to obtain the vaccine if they are at a group recommended for vaccination (e.g. health workers), and does not advise cessation of breastfeeding following receipt of the vaccine ^4^. The Academy of Breastfeeding Medicine (ABM) states that there is little plausible risk that vaccine lipid particles would enter the blood stream and be present in breast tissue, and that nanoparticles or mRNA would be transferred to milk ^5^. However, regardless of the low predicted risk of harm to the baby, some mothers have declined vaccination, chosen to “pump and dump” breast milk for up to 72 hours after the vaccine, or decided to stop breastfeeding due to the lack of solid evidence about the effect of the mRNA vaccine on human milk.

We herein report that analysis of milk samples collected from six individuals within 24 hours after mRNA vaccination against COVID-19, and of six serial milk samples collected 4, 8, 22, 28, 33, and 48 hours post vaccine showed no evidence of vaccine-related mRNA in breast milk.

## Methods

The University of California San Francisco (UCSF) institutional review board approved the study (20-32077). Informed consent was obtained from all study participants.

Human breast milk samples were collected fresh or frozen (immediately after milk was pumped). Total RNA was isolated from milk components (cells, milk supernatant and/or fat layer) using the RNeasy Mini Kit (Qiagen) according to manufacturer’ s protocol. We performed RT-qPCR in triplicate using specific primers (supplementary materials) targeting the vaccines mRNA for SARS-CoV-2 spike protein. mRNA-1273 (Moderna) vaccine was spiked into pre-vaccine milk sample before RNA isolation and served as a positive control for this assay. Pre-vaccine samples served as negative controls.

## Results

Post-vaccine human milk samples were collected from six individuals 4-48 hours after administration, 5 vaccinated with BNT162b2 (Pfizer) and 1 individual with mRNA-1273 (Moderna) vaccine (**Table 1**). We first optimized our RT-qPCR by isolating the residual vaccine mRNA from vials, showing that our assay is capable to detect up to 1.5 pico grams of the mRNA-1273 vaccine cDNA and up to 0.195 pico grams of the BNT162b2 vaccine (**Figure 1A**). We next used pre-vaccine milk samples and spiked-in the mRNA-1273 vaccine (12 and 0.12ng/ul vaccine mRNA). RNA was extracted from the supernatant and fat layer of these spiked-in milk samples. We were able to detect the spiked-in vaccine mRNA in these samples (**Figure 1B**), with higher levels of vaccine mRNA in fat layer fraction (**Figure 1B**). We next analyzed 12 post-vaccine samples (4-48 hours post vaccine, **Table 1**) and found that none of the samples from vaccinated lactating mothers showed detectable levels of vaccine mRNA in milk fat layer or milk supernatant at any time point (7 samples from 24h post vaccine are shown in **Figure 1B**).

**Table 1:** Samples analyzed by qPCR analysis

| Participant number | Vaccine | Time point | Collection Method |  | Milk Fraction |  |  |
| --- | --- | --- | --- | --- | --- | --- | --- |
|  |  |  | Fresh | Frozen | Supernatant | Fat | Cells |
| 1 | BNT162b2 | Pre-vaccine |  | X | X |  |  |
|  |  | 24h postvaccine 1 |  | X | X | X |  |
| 2 | BNT162b2 | Pre-vaccine |  | X | X | X |  |
|  |  | 24h postvaccine 1 |  | X | X | X |  |
|  |  | 24h postvaccine 2 | X |  | X | X |  |
| 3 | BNT162b2 | 24h postvaccine 1 | X |  | X | X |  |
| 4 | mRNA-1273 | 8h postvaccine 1 |  | X | X | X |  |
|  |  | <24h postvaccine 1 |  | X | X | X |  |
|  |  | 28h postvaccine 1 |  | X | X | X |  |
|  |  | 33h postvaccine 1 |  | X | X | X |  |
|  |  | 48h postvaccine 1 |  | X | X | X |  |
|  |  | 4h post vaccine 2 |  | X | X | X |  |
| 5 | BNT162b2 | 24h postvaccine 1 | X |  | X | X |  |
| 6 | BNT162b2 | Pre-vaccine |  | X | X | X | X |
|  |  | 24h postvaccine 1 | X |  | X | X | X |

**Figure 1.**
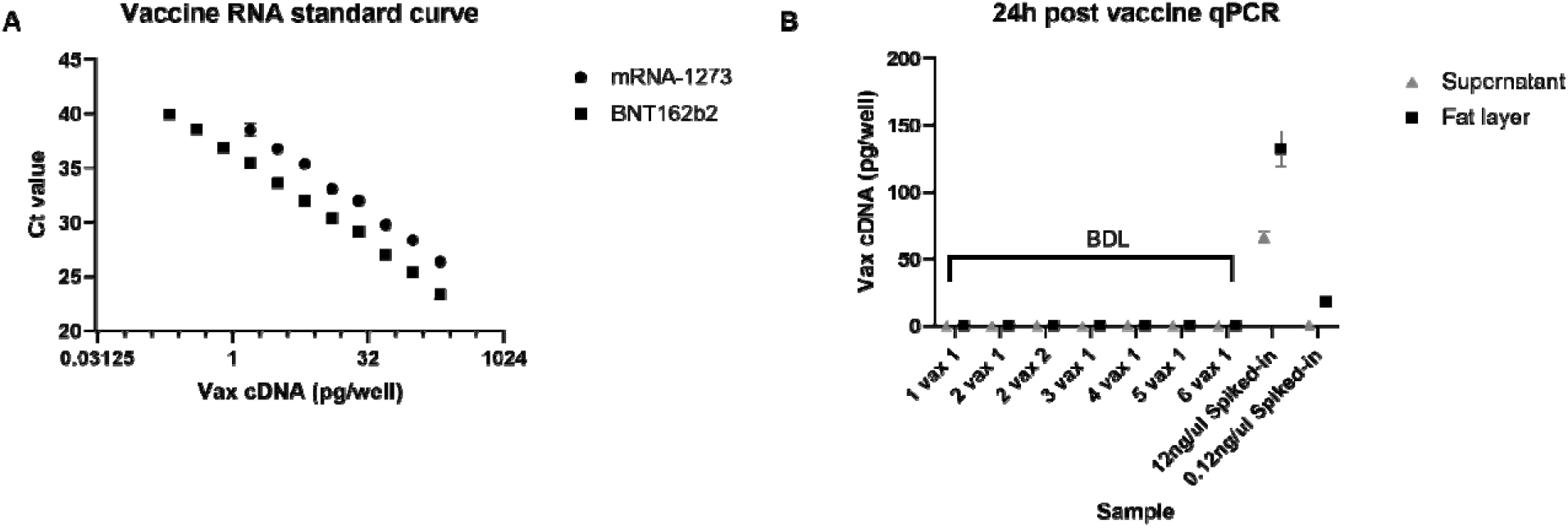
qPCR analysis of post vaccine milk samples. A) standard curves for mRNA-1273 and BNT162b2 vaccines. B) mRNA concentrations of 24h post vaccine samples and spiked-in samples were calculated based on equations from standard curves. Samples names stands for participant number and if sample was collected after first (1) or second (2) vaccine (vax). Below detectable levels (BDL). Mean values ± SD are shown.

## Conclusion

We show here that the mRNA from anti-COVID vaccines is not detected in human breast milk samples collected 4-48 hours post-vaccine. These results strengthen the recommendation of ABM and WHO that lactating individuals who receive the anti-COVID-19 mRNA-based vaccine should continue to breastfeed their infants uninterrupted. Clinical data from larger populations need to be collected and analyzed to better estimate the effect of these vaccines on lactation outcomes.

## Data Availability

All data from this manuscript will be available.

## Acknowledgment

We would like to acknowledge the milk donors that volunteered to this study. In addition, we would like to acknowledge Kenneth Scott and Hannah Jang MSN, PhD for their contribution to this study.

