## supplementary materials for "COVID-19 mRNA vaccine is not detected in human milk"

### Supplementary material

*Milk samples-* Fresh human milk samples were self-collected by participants 24h after vaccine administration. If not collected immediately, mothers were instructed to freeze milk as soon as possible after milk pumping. Samples were kept on ice until arriving in the lab for processing (up to 2 hours after milk pumping). Five ml of whole breast milk was mixed in a 1:1 ration with RNAlater (catalog number R0901; Sigma-Aldrich), aliquoted and frozen at -80°C until analyzed (Breast milk in RNAlater was treated as the frozen sample below). The remaining milk was centrifuged for 20 min at 800g at 4°C. Approximately 200ul of fat layer was collected into new Eppendorf tubes and was mixed with 600 µL of RLT buffer (RNeasy Mini Kit; Qiagen) and kept in -80°C until analyzed. Supernatant was removed and cells pellets were resuspended in 350 µL of RLT buffer (cells fraction).

Frozen milk samples were thawed on ice and were mixed gently by pipetting up and down. 1 ml milk was mixed with RNAlater in 1:1 ratio immediately after thawing. Samples were centrifuged for 1 min 10000g at 4°C to separate milk fat from supernatant. Milk fat was transformed to a new tube and was mixed with 600 µL RLT buffer immediately. Two hundred µL of supernatant were mixed with 600 µL RLT buffer. Samples in RLT buffer were used for RNA isolation. Fat layer samples in RLT buffer were centrifuged for 1 min 10000g at 4°C and fat was removed from samples before RNA isolation.

*BNT162b2 (Pfizer) and mRNA-1273 (Moderna) mRNA PCR* - RNA was isolated from samples using the RNeasy Mini Kit (Qiagen) according to manufacturer's protocol. RNA concentration was measured using nanodrop and samples that had >10ng/µl total RNA were used for RT reaction. 150-500ng RNA was transcribed into cDNA using qScript cDNA synthesis kit (Quantabio) according to the manufacturer's protocol. Primers were design to detect the vaccines mRNA, and BNT162b2 (Pfizer) and mRNA-1273 (Moderna) commercial vaccine was used to determine primers specificity and sensitivity. Forward primer: AACGCCACCAACGTGGTCATC. Reverse primer: GTTGTTGGCGCTGCTGTACAC. For positive control, 30 µL (200ng/µL) of mRNA-1273 (Moderna) were spiked-in to 500 µL of whole milk (12ng/µL). This sample was diluted in 1:100 in whole milk to create the 0.12ng/µL sample. The spiked in samples were mixed with RNAlater in 1:1 ration and treated as described above for RNA isolation from milk samples.

QuantaStudio 6 Flex (Applied Biosystems) instrument and SsoFast EvaGreen supermix (Bio-Rad) were used for PCR reaction: 30 second 95°C followed by 45 cycles of 5 second 95°C and 20 seconds 60°C.

All samples were run in triplicate as 20 µL reactions, containing 10 ng cDNA. Ct values ≥40 were interpreted as a negative result (BDL, below detectable levels). Threshold was set based on negative controls of pre-vaccine samples and NTC. For vaccines cDNA standard curves, 100pg/µL vaccine mRNA (as cDNA) sample was used for serial dilution in 1:2 ratio, up to 0.0975 pg/µL. Two µL of these diluted samples were used in each well to create standard curves.

*Calculation of samples concentration*- Vaccine cDNA was used to create standard curves. Log values of pg/wells were plotted on X axis and Ct values on Y axis. The mRNA-1273 vaccine equation was $y = -6.522x + 38.437$, and *BNT162b2* vaccine equation was $y = -4.8558x + 38.898$. Ct values of samples were used to calculate pg vaccine cDNA in each well.
